# Cerebri: a web-app to reduce door-to-treatment decision time and improve guideline adherence in acute ischaemic stroke

**DOI:** 10.1101/2025.08.01.25332640

**Authors:** A. Bonura, G. Musotto, S.S. Rossi, F. Santoro, P. Pillitteri, M. Sferruzzi, V. Di Lazzaro, F. Pilato

## Abstract

**Background:** Clinical Decision Support Systems (CDSS) hold promise for improving adherence to guidelines and reducing decision-making time in acute ischemic stroke management. We developed a stroke-specific CDSS smartphone app termed Cerebri, and assessed its efficacy, in reducing imaging-to-decision time (IDT) and enhancing guideline adherence in a real-world clinical setting.

**Methods:** We conducted a prospective, single-center, cluster-randomized pilot study at the Emergency Department of Campus Bio-Medico University Hospital in Rome, Italy, from December 2024 to March 2025. Eligible participants were adult patients (≥18 years) with suspected ischemic stroke presenting within 24 hours of symptom onset. Randomization occurred by day, assigning patients to either the Cerebri group (app providing clinical decision support plus a timer function) or the control group (timer function only, without decision support). Primary outcomes were IDT (imaging to decision time: the time from imaging completion to therapeutic decision) and adherence to European Stroke Organisation guidelines. Multivariable linear and logistic regression analyses adjusted for age, NIHSS, wake-up stroke status, revascularization therapies, hemorrhagic stroke, and stroke mimics were performed.

**Results:** Fifty patients (mean age 73.1±13.3 years, 64% female) were enrolled (25 Cerebri, 25 control). Patients in Cerebri group had significantly reduced median imaging to decision time (6 vs. 22 minutes; adjusted difference: −18.9 minutes, 95% CI −32.4 to −5.2, p=0.006). Overall guideline adherence was significantly higher in the Cerebri group (96% vs. 73.9%; adjusted OR 11.6, 95% CI 1.12–121.16, p=0.040). Therapeutic guideline adherence was 100% in the Cerebri group vs. 86.9% in control (p=0.062).

**Conclusions:** Cerebri significantly improved decision-making speed and adherence to stroke guidelines in the hyperacute management of ischemic stroke. The app demonstrated feasibility and efficacy even in non-tertiary stroke centers, suggesting its potential for broader implementation to enhance stroke care quality and equity.

## 1. INTRODUCTION

Stroke is one of the leading causes of death and disability worldwide, with more than 12 million new cases annually and healthcare costs exceeding 60 billion EUR in Europe, and 45.5 billion USD in USA^1–3^.

Timely intervention in acute stroke management is crucial, as approximately 1.9 million neurons are lost every minute delay in revascularization, and every 15 minutes delay leads to an increased risk of long-term disability, in-hospital mortality, and intraparenchymal hemorrhage, clearly illustrating the “time is brain” paradigm^4–6^.

This clinical burden translates into a substantial economic impact: each 10-minute delay in endovascular thrombectomy (EVT) has been associated with a loss of approximately 0.24 disability-adjusted life years (DALYs), equivalent to about 2.9 months of healthy life per patient, and a median net monetary loss of €309 per minute of delay based on a willingness-to-pay threshold of €80,000 per quality-adjusted life year (QALY)^7,8^.

Recent updates to the European guidelines for acute ischemic stroke management, including extended treatment windows and advanced imaging criteria, have substantially increased the complexity of decision-making algorithms^9–11^. While these updates aim to optimize clinical outcomes, they paradoxically introduce additional challenges for emergency clinicians, potentially leading to delays and suboptimal therapeutic decisions^12^. The heightened complexity demands more specialized training and expertise, further widening disparities between Comprehensive Stroke Centers (CSCs) and Primary Stroke Centers (PSCs)^13^. Consequently, strategies to standardize stroke care become essential for bridging these gaps and ensuring more equitable patient outcomes.

Computational tools are increasingly transforming medical practice, playing a critical role in various aspects of healthcare, from artificial intelligence-based diagnostic algorithms to sophisticated simulations of clinical scenarios^14–17^. In particular, Clinical Decision Support Systems (CDSS) represent a promising application of computational support, and have proven effective in enhancing the management of complex acute conditions by making guidelines interactive and streamlining clinical decision-making processes^18–20^. Different applications have been proposed in the stroke field, such as JOIN or FAST-ED that have demonstrated significant reductions in stroke intervention times and improved early detection of large vessel occlusions^21–24^. Despite these advances, there remains a notable scarcity of CDSS specifically designed to assist clinicians in the hyperacute phase of ischemic stroke^19^.

In response to these challenges, we developed Cerebri, an in-house mobile Clinical Decision Support System (CDSS) tailored to hyperacute ischemic stroke management. The app embeds the latest European Stroke Organisation (ESO) guidelines into an interactive, cascading “if–then” algorithm: each clinician input automatically triggers the next context-specific module, ultimately generating patient-specific recommendations for both imaging and treatment (Figure 1).

**Figure 1.**
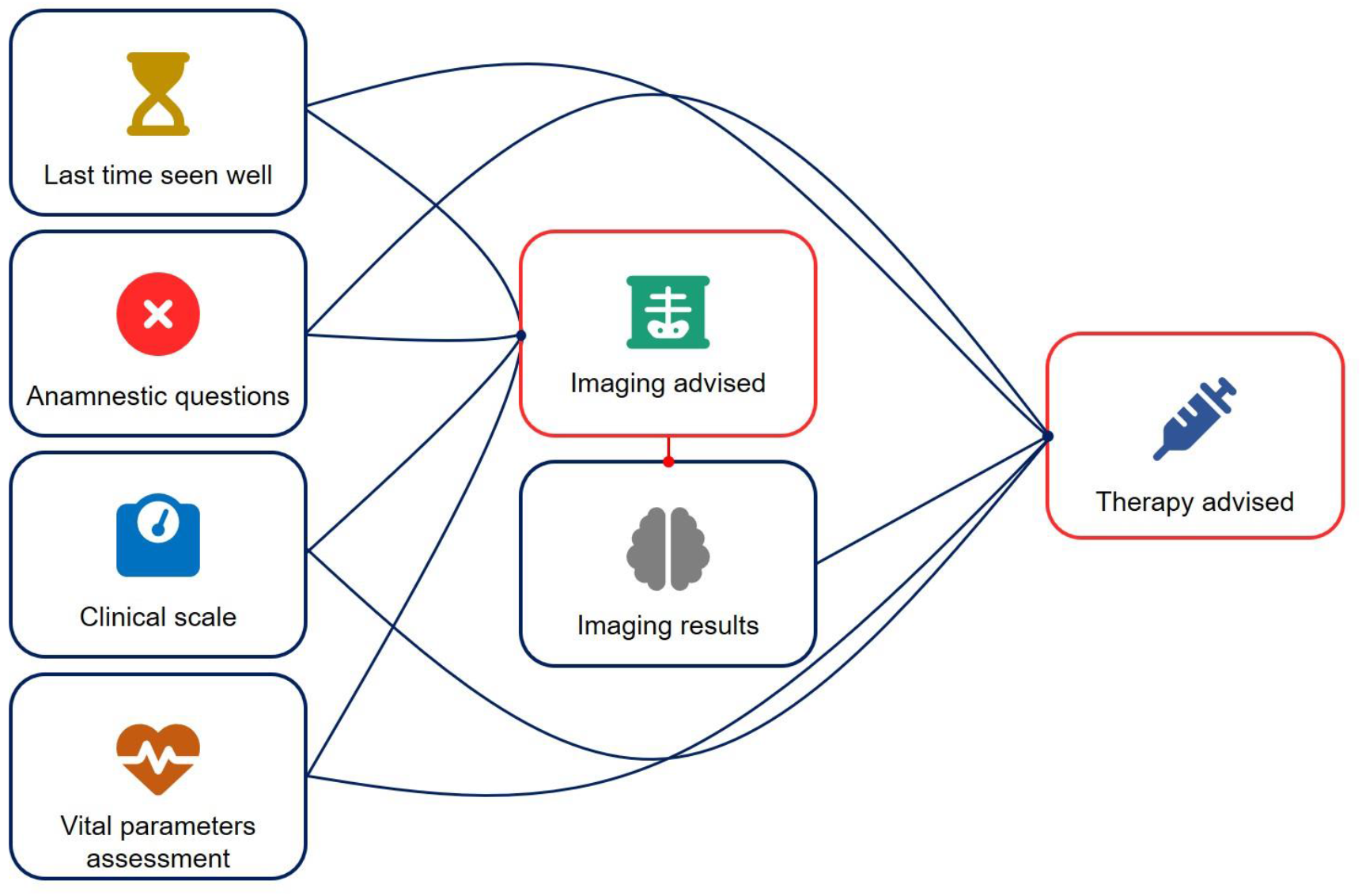
App workflow diagram. Blue boxes represent input functions; red boxes represent outputs. Clinical data such as last time seen weel, anamnestic questions, NIHSS, mRS, laboratory data, and vital parameters are first entered into the app, which then determines the most appropriate imaging modality as output. Once imaging results are available the user can enter it as a new input, the app combines them with the previously submitted clinical data to calculate the most suitable therapeutic strategy. Icons obtained from fontawesome.com (free license).

**Figure 2.**
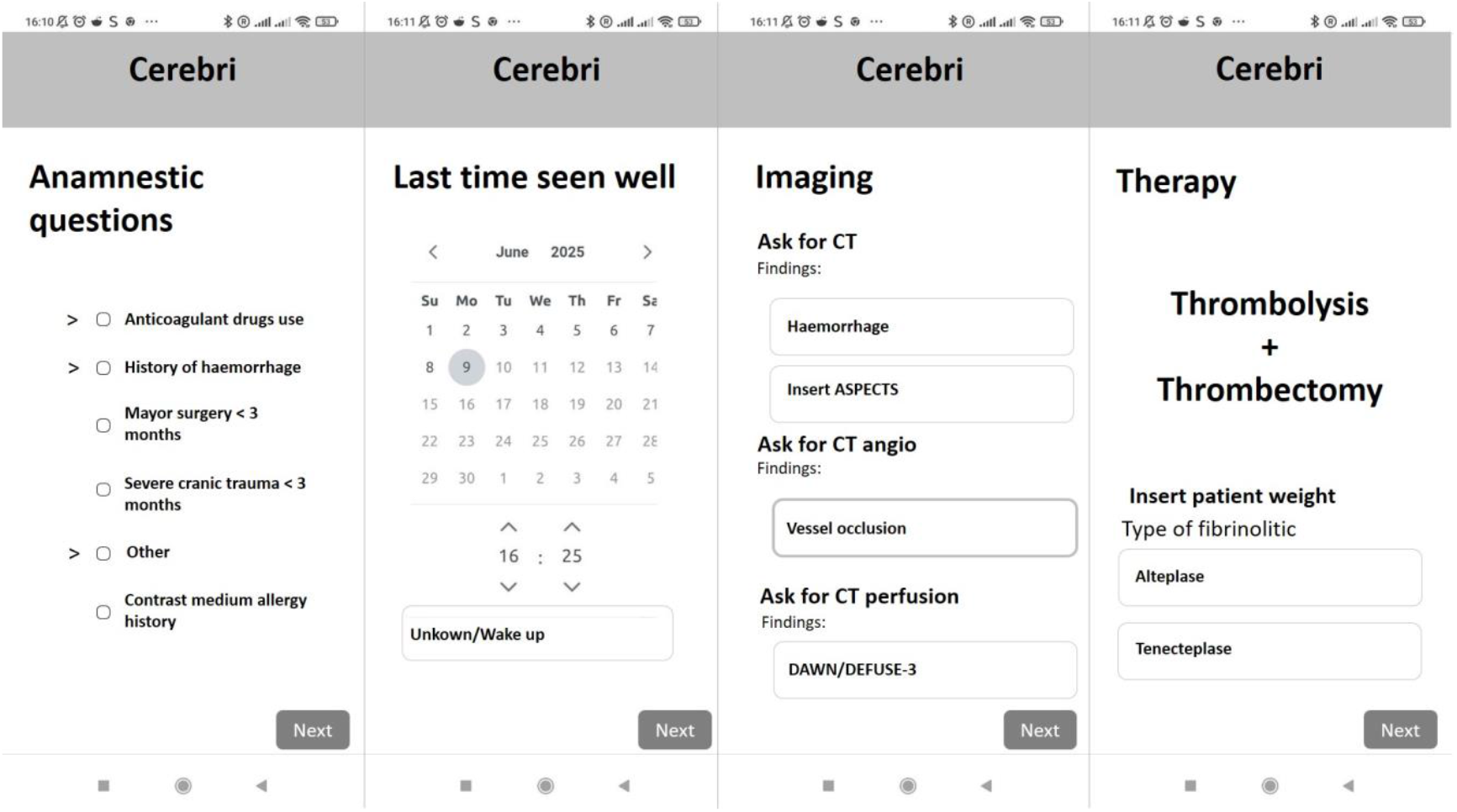
Cerebri mock-up. The app guides the clinician through the most relevant clinical questions to determine the appropriate therapy and identify the optimal imaging modality. Built-in calculators are available for NIHSS, mRS, and dosing of Alteplase and Tenecteplase. Additionally, the interface includes direct links to current guidelines and key studies (e.g., DAWN/DEFUSE-3, EXTEND criteria). These images are illustrative mock-ups and do not represent actual screenshots of the app.

**Figure 3.**
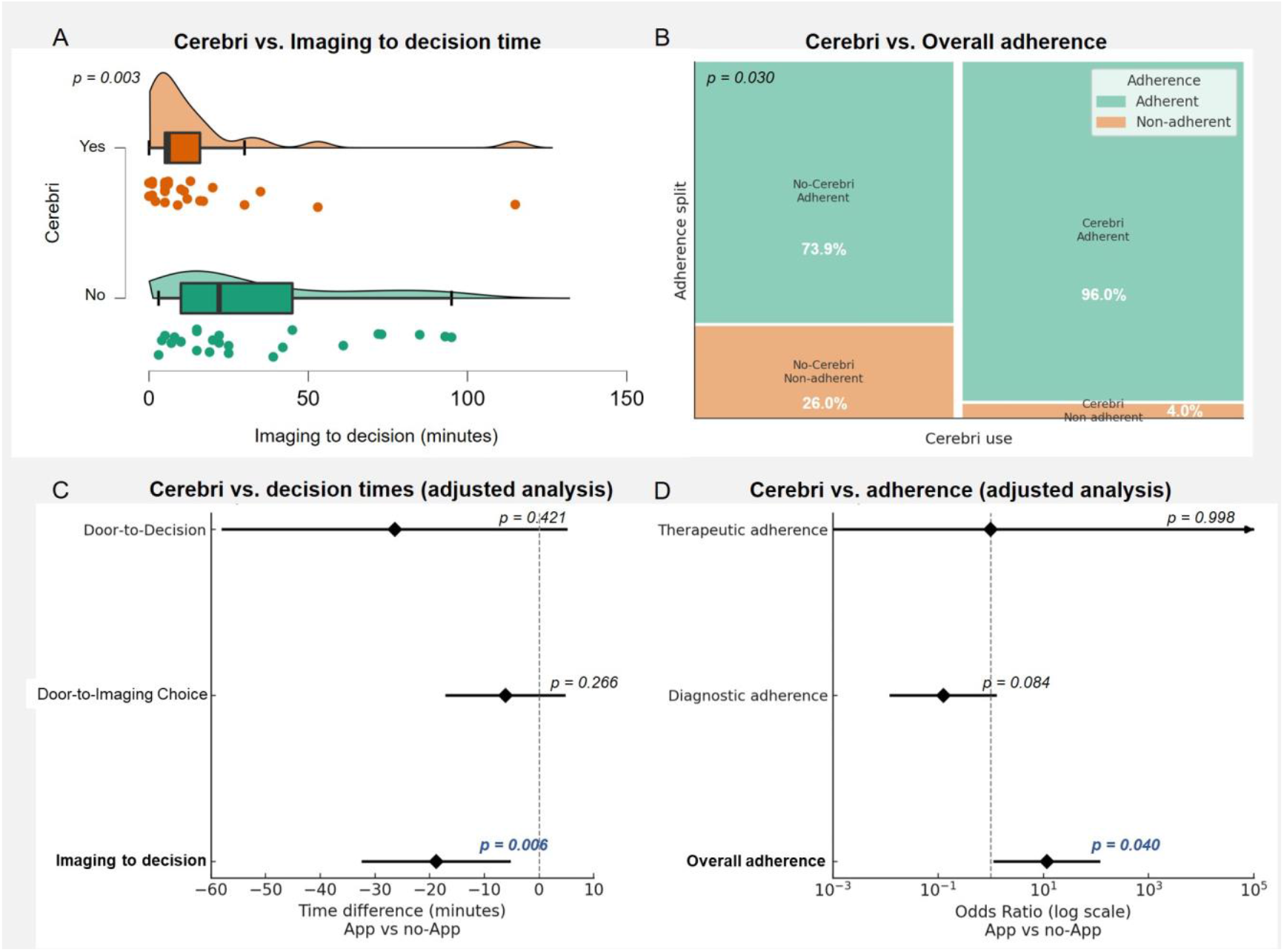
Impact of Cerebri on decision-making times and guidelines adherence. A) Violin plot of distribution of imaging-to-decision times (minutes) on days with Cerebri (orange) versus without Cerebri (green). Beans depict the density; squares mark the median and whiskers the IQR. Mann Whitney test: p = 0.003. B) Mosaic plot of overall adherence to ESO recommendations. Rectangle areas are proportional to patient counts. Adherence rises to 96 % with Cerebri compared with 74 % without the app (p = 0.030). C) Adjusted linear effects of Cerebri on three time-to-treatment endpoints. Bars show 95 % CIs; negative values favour the app. Only the imaging-to-decision interval is significantly shortened (–18.9 min; p = 0.006). D) Adjusted logistic models for adherence outcomes (log-scale odds ratios). Cerebri significantly increases overall adherence (OR 11.6; p = 0.040). Effects on diagnostic adherence (borderline, p = 0.084) and therapeutic adherence (upper CI unbounded, p = 0.998) are not significant.

This study evaluates whether Cerebri shortens decision-making times and improves adherence to guideline-based diagnostic and therapeutic pathways, thereby making evidence-based stroke care more accessible and easier to apply at the bedside.

## 2. METHODS

### Study Design

We conducted a prospective, single-centre, cluster-randomised, open-label, two-arm pilot trial in the Emergency Department (Stroke Spoke Centre) of Campus Bio-Medico University Hospital, Rome, Italy. Consecutive patients were screened and enrolled between 1 December 2024 and 30 March 2025. The study was reported in accordance with the Consolidated Standards of Reporting Trials (CONSORT) statement and its accompanying checklist^25^.

### Inclusion criteria

Adults (≥ 18 years) with suspected anterior- or posterior-circulation acute ischaemic stroke presenting within 24 h of last-known-well are eligible. Major exclusion criteria are patients with a suspected transient ischaemic attack (TIA) and an NIHSS score of 0 at emergency department admission, participation in another interventional trial, lack of availability of imaging modalities required for decision-making (e.g., CT, CTA, or MRI), unavailability of thrombolytic treatment or endovascular suite at the time of evaluation.

### Randomization and Allocation

Study days within the recruitment window were pre-randomised in a 1:1 ratio to either the Cerebri intervention or the standard-care control arm using a computer-generated sequence (simple random sampling without replacement). The allocation schedule was created by an independent statistician and stored on a secure server; the on-call stroke team was informed of the day’s assignment at 07:00 each morning. All eligible patients presenting on an “intervention day” were managed with Cerebri, whereas those presenting on a “control day” received usual care based on paper/electronic guidelines and they used the app only in timer function. Because allocation occurred at the day level, outcome assessors and data analysts remained blinded to treatment conditions.

### Sample Size Calculation

This pilot study was designed to assess feasibility and preliminary efficacy of the Cerebri application. Randomisation occurred at the level of calendar day (cluster-randomised design), with each day allocated to either the Cerebri or control group. From preliminary observation, an average of 2.1 patients presented per day.

Assuming an intracluster correlation coefficient (ICC) of 0.05 and a mean cluster size of 2.1, the design effect (DE) is estimated as DE = 1 + (m − 1) × ICC ≈ 1.05. The ICC value of 0.05 was selected in line with recommendations for pilot trials involving small cluster sizes, such as those by Eldridge et al. (2015), where modest intra-cluster correlation is anticipated due to shared personnel or logistics^26^. In our setting, potential sources of intra-day correlation include consistent clinical staffing, similar workflows, and environmental constraints, all of which might mildly influence decision-making behavior.

To estimate the number of patients required, we targeted an 80% power to detect a between-group difference in imaging-to-decision time (IDT) of 15 minutes, assuming a standard deviation of 15–20 minutes based on prior studies of acute stroke workflows. This delta was chosen as a conservative yet clinically meaningful threshold, given that even small improvements in treatment timing can yield substantial outcome and cost benefits in hyperacute stroke care. Under individual-level randomisation, this would require approximately 46–48 patients. To account for clustering by day, this effective sample size was inflated using the design effect, yielding a total of approximately 50 patients required. The design effect ensures proper adjustment for the small degree of within-day dependency introduced by the cluster randomisation scheme.

### Data Collection and Outcomes

We collected demographic data (age and gender), clinical data (NIHSS at admission, pre-stroke mRS, haemorrhagic stroke, stroke mimic), and administered revascularization therapies (rTPA, EVT, or both). Additionally, we recorded various time intervals from patient arrival in the Emergency Department to imaging decision (Door to imaging choice time), therapeutic decision (door to decision time and imaging to decision time), and the administration of therapies (door to needle and door to groin times). Time intervals were recorded using Cerebri’s internal timer in both study arms: in the intervention group, the app provided real-time clinical suggestions alongside automatic time tracking, while in the control group, only the timer function was available, with no decision-support features enabled.

*Primary outcomes* include (1) imaging to decision time (IDT), defined as the interval between completion of first neuro-imaging and documentation of the therapeutic decision in the electronic medical record, and (2) guideline adherence.

IDT was selected as a primary endpoint because it directly captures the decision-making latency independent of pre-imaging delays or the type of imaging performed. In our pilot cohort, imaging modality was largely homogeneous, with most patients undergoing non-contrast CT and CT angiography ± perfusion, which minimized its direct influence on therapeutic decision time. However, we observed substantial variability in the timing of imaging acquisition itself—due to factors such as scanner availability, emergency department crowding, and type of imaging requested—which could introduce potential bias into time-based outcome measurements.

Adherence with guideline-based decision-making was assessed separately for imaging selection and therapeutic choice. Each case was independently reviewed by two blinded senior neurologists (P.F, D.L.V.) who evaluated whether the imaging modality selected and the final therapeutic decision (e.g., thrombolysis, transfer for EVT, or no treatment) aligned with the most recent ESO guidelines. Discrepancies were resolved through consensus. A binary score (compliant vs. non-compliant) was assigned for both diagnostic and therapeutic decisions, and overall adherence was defined as concordance with both elements. For this assessment, both clinical records and the data automatically recorded by the app—detailing each decision made during the workflow—were reviewed. Cases with missing data or incomplete documentation were excluded from the adherence analysis.

Secondary outcomes include classical metrics such as door-to-needle time (DNT), along with additional derived intervals: door-to-imaging-choice, and door-to-decision times. Door-to-groin time was not included as an endpoint, as our spoke centre does not perform thrombectomy onsite; EVT is performed after patient transfer, making these timings dependent on external logistics and not directly modifiable by the intervention.

Moreover, we examined the learning effect specifically in the population not exposed to the app, considering that the cluster-randomized design allowed the same neurologist to use the app on previous days and potentially retain familiarity with the decision-making process. This approach enabled us to assess whether clinical improvements extended beyond direct usage to influence routine care.

### Statistical Analysis

Continuous variables were presented as means with standard deviations if normally distributed, and as medians with interquartile ranges (IQRs) if non-normally distributed. Normality was assessed using the Shapiro–Wilk test and visual inspection of histograms. Categorical variables were reported as absolute counts and percentages.

Intracluster correlation coefficients (ICCs) were computed for both continuous and categorical endpoints to evaluate the extent of clustering by day. For continuous outcomes, ICCs were estimated using one-way random-effects ANOVA models. For categorical outcomes, the ICC was approximated using the Fleiss–Cuzick estimator based on variance components from logistic regression. In all cases, the ICC was found to be <0.01, indicating negligible within-day correlation.

Accordingly, continuous outcomes (e.g., IDT, door-to-needle time) were compared between groups using unpaired two-sample t-tests or Mann–Whitney U tests, depending on data normality. Categorical variables (e.g., guideline adherence, symptomatic intracranial haemorrhage) were analysed using chi-squared or Fisher’s exact tests as appropriate. Additionally, we performed multivariable analyses to adjust for potential confounders. For the continuous outcomes, a linear regression model was constructed including age, baseline NIHSS, administration and type of revascularization therapy (thrombolysis or EVT), presence of haemorrhagic stroke, suspected stroke mimic, and wake-up stroke status. For the binary outcome of guideline adherence, a logistic regression model was used adjusting for age, NIHSS, and type of revascularization therapy. All tests were two-sided with a significance threshold of p < 0.05.

To investigate whether cumulative exposure to the Cerebri app improved decision-making performance even in subsequent cases where the app was not used, we conducted two separate models: a linear mixed-effects model for continuous process variables (IDT, DNT, Door to imaging choice time, Door to decision time) and a logistic regression model for the binary outcome variable (Overall adherence, Diagnostic adherence, Therapeutic adherence). In both cases, the day of patient admission to the Emergency Department was used as a proxy for cumulative team exposure to the app. All models adjusted for relevant clinical covariates: revascularization therapy, hemorrhagic stroke, stroke mimics, wake up stroke, extendend windows stroke management (>4.5 h for thrombolysis and > 6 h for thrombectomy), and baseline stroke severity, excluding those with no variability in the subset analyzed. Model estimates were obtained using maximum likelihood estimation (REML = FALSE for linear models), and statistical significance was assessed using Satterthwaite’s method (for mixed models) or Wald tests (for logistic regression). Analyses were performed using R 4.3.1 with standard statistical libraries. The plots were created with Jasp (v 0.16.4) and matplotlib library (Python v 3.14).

### Ethics

The study protocol has been reviewed and approved by the Institutional Review Board of Campus Bio-Medico University of Rome (Protocol ID: **PAR 18.23**.). Informed consent was obtained from each participant or, when applicable, from their legally authorised representative, in accordance with the Declaration of Helsinki and EU GDPR regulations.

## 3. Results

The mean age was 73.1 ± 13.3 years, with 64% female participants. Of the 50 patients included, 43 were diagnosed with ischaemic stroke, 5 with haemorrhagic stroke (10%), and 3 with stroke mimics (2 epileptic seizures, 1 tumour). The median NIHSS at admission was 8 (IQR 4.2–12), with no significant difference between groups. A total of 17 patients (34%) received intravenous thrombolysis and 14 (28%) underwent EVT; the latter occurred predominantly in the control arm (11 patients vs. 3).

Regarding revascularisation timing, the primary endpoint, imaging-to-decision time (IDT), showed a substantial and statistically significant reduction in the Cerebri group (6 [IQR 5–16] minutes vs. 22 [10–45] minutes, p = 0.003). Adjusted linear regression confirmed a reduction of 18.9 minutes in the intervention group (t= −2.91, 95% CI −32.4 to −5.2, p = 0.006).

No significant difference was observed in door-to-needle times between groups, with a median of 63 minutes overall (IQR 40–80), 63.5 minutes in the Cerebri group (IQR 34.7–74.7), and 63 minutes in the control group (IQR 45–85) (p = 0.413).

Among others timing parameters, the median door-to-imaging-choice time was 8.0 minutes (IQR 5– 13), with 7 minutes in the Cerebri group (IQR 5–12) and 9 minutes in the control group (IQR 3–13) (p = 0.938), showing no significant difference. The median door-to-therapy-decision time was 47.0 minutes (IQR 27.7–86.2), with 40 minutes in the Cerebri group (IQR 25–65) and 70 minutes in the control group (IQR 32–97), showing an almost-significant trend toward improvement with Cerebri (p = 0.061).

Guideline adherence was achieved in 96% of Cerebri cases (24 patients) versus 73.9% (17 out of 23) in the control group (p = 0.030). The adjusted logistic regression model demonstrated a statistically significant improvement in overall adherence in the Cerebri group compared to control (z = 2.056, OR = 11.6; 95% CI: 1.12–121.16, p = 0.040). Imaging selection complied with guidelines in 96% of cases in the Cerebri arm versus 80% in the control arm (p = 0.180). In the control group, perfusion imaging was omitted in four cases despite being indicated, whereas in the Cerebri group, one patient underwent CT perfusion despite having both a high pre-stroke mRS (indicating a contraindication for thrombectomy) and contraindications to rTPA due to anticoagulant therapy. Regarding therapeutic decision-making, appropriate treatment was administered in 100% of Cerebri cases compared to 86.9% in the control group (p = 0.062); notably, three patients in the control arm did not receive rTPA despite the absence of contraindications.

The learning effect analysis did not show a learning curve in the without-app arm both for IDT and for guidelines’ adherence (p=0.891, p=0.451 respectively)

## 4. Discussion

Stroke is one of the medical emergencies with the highest clinical and socioeconomic impact. The growing complexity also related to continuous updating and urgency of acute stroke management necessitate specialized expertise and dedicated infrastructure, resulting in substantial disparities between tertiary and non-tertiary hospitals. Evidence indicates that specialized centers achieve superior outcomes, demonstrating higher rates of timely reperfusion therapies—both thrombectomy and administration of rTPA—, best adherence to stroke guidelines, and better functional outcomes compared to smaller hospitals^13,27^. Cerebri aims to mitigate these disparities by standardizing stroke care, making guidelines easier to read and interpret, thereby facilitating consistent and guideline-based clinical decisions. The effectiveness of this approach has been quantitatively demonstrated by a significant reduction in decision-making times and enhanced adherence rates.

In this study Cerebri arm showed an 18-minute reduction in imaging-to-decision time, corresponding to the preservation of approximately 34.2 million neurons, 252 billion synapses, and 216 kilometers of myelinated fibers^5^. According to Sheth et al. such improvements can potentially reduce disability in approximately 41 out of every 1,000 patients^28^. However, this result is tempered by the lack of improvement observed in the door-to-needle time. On one hand, this finding underscores persistent issues in our center with imaging acquisition times, which indeed appear to be a major contributor to the unchanged door-to-needle intervals. On the other hand, the limited sample size of patients receiving thrombolysis (8 in the Cerebri group and 9 in the control group) may also have contributed to the absence of statistically significant differences for this outcome.

The improved adherence rates observed with Cerebri support the role of CDSS tools in bridging the gap between clinical evidence and real-world practice. In our study, overall guideline adherence improved from 73.9% in the control group to 95% (+21.1%) in the Cerebri group, with therapeutic decision adherence rising to 100% versus 86.9% in the control arm (+13.1%). These improvements are particularly relevant in fast-paced or resource-limited environments, where rapid decision-making and guideline interpretation may be more challenging. Furthermore, several studies have shown that adherence to stroke management guidelines is significantly associated with improved patient outcomes, including reduced mortality and long-term disability. For example, the “Get With The Guidelines–Stroke” program by the American Heart Association demonstrated that consistent implementation of evidence-based protocols leads to lower complication rates and better functional recovery.^29^

Another important point is the potential of Cerebri to enhance and accelerate clinicians’ understanding and assimilation of stroke management guidelines. Although the learning effect analysis did not yield statistically significant results, likely due to the limited number of patients and the short duration of the study period, it suggests a potential educational benefit associated with the repeated use of the app. This finding supports the idea that CDSS tools like Cerebri may not only aid real-time decisions but also contribute to the internalization and reinforcement of clinical guidelines, promoting more consistent care over time.

Finally, the successful deployment of Cerebri in a spoke center demonstrates its feasibility in non-tertiary clinical environments. Unlike comprehensive stroke centers, Spoke hospitals often lack round-the-clock access to neurointerventionists and advanced imaging interpretation. The context in which Cerebri was implemented reflects these constraints, reinforcing the applicability of its benefits to a broad spectrum of real-world settings.

## 5. Conclusions

In conclusion, Cerebri proved to be a valuable tool in the hyperacute management of ischemic stroke, demonstrating improvements in decision-making times and adherence to clinical guidelines. Furthermore, its successful implementation in non-tertiary centers underscores its potential to reduce systemic disparities in care access and quality. Taken together, these findings support the integration of Cerebri not only as a decision aid but also as an educational, organizational, and economic lever to promote equitable, standardized, and high-quality stroke care across diverse healthcare settings.

## Data Availability

Data will be available under justified request

## 6. Limitations

This study has some limitations that should be acknowledged when interpreting the findings. First, the single-center design may affect the generalizability of the results to other institutions with different workflows, staffing, or resources. Nonetheless, the study was conducted in a representative spoke center—typical of many real-world stroke care environments—thereby enhancing the applicability of the results in settings with similar constraints. Furthermore, the use of IDT as a primary outcome helped mitigate potential bias arising from differences in resources and organizational structures between hospitals.

Another limitation was the small number and an unbalanced distribution of endovascular thrombectomy procedures, with 11 patients undergoing EVT in the control group versus only 3 in the Cerebri group. This disparity limits conclusions regarding the app’s specific impact on EVT timing. However, appropriate regression models were used to adjust for potential confounders.

We are aware that a potential learning effect may have occurred. Repeated use of the decision-making workflow and familiarity with the app’s logic could have improved clinical performance over time, regardless of group allocation. Nevertheless, our analysis did not show a statistical significative learning effect, this could be due to the high number of neurologists involved in on-call rotation (12 neurologists) at Campus Bio Medico University Hospital, and the fact that the app have been used just a few times by the same practicioner.

Finally, the study did not include long-term clinical outcomes such as functional independence at 90 days, focusing instead on process-related endpoints like time metrics and guideline adherence. This is consistent with the exploratory nature of the study, which aimed to assess the tool’s impact on hyperacute phase decision-making rather than definitive clinical outcomes.

---

**Table 1.** Data collected. Significative differences between the two arms are highlighted in bold. ^*^The Cerebri group includes only 3 patients in this subgroup.

| Variable | Value |  |  |  |
| --- | --- | --- | --- | --- |
|  | Total | Cerebri | Control | p-value |
| Age | 73.1 +- 13.3 | 69.5 +- 14.9 | 77.1 +- 10.2 | 0.061 |
| Female | 32 (64%) | 14 (56%) | 18 (72%) | 0.239 |
| NIHSS at admission | 8 (IQT= 4.2 – 12) | 7 (IQT = 4.5 -10) | 8 (IQT = 4.5 – 12.5) | 0.639 |
| mRS pre-stroke | 2 (IQT = 0 - 3) | 2 (IQT = 0 - 3) | 2 (IQT = 1 - 4) | 0.890 |
| Haemorrhagic stroke | 5 (10%) | 5 (20%) | 0 | **0.018** |
| Stroke mimic | 3 (6%) | 3 (12%) | 0 | 0.074 |
| Wakeup stroke | 19 (38.8%) | 11 (44%) | 8 (36%) | 0.444 |
| Extended windows stroke | 15 (30%) | 7 (14%) | 8 (16%) | 0.857 |
| rTPA | 17 (34%) | 8 (36 %) | 9 (36 %) | 0.765 |
| EVT | 14 (28%) | 3 (12 %) | 11 (44 %) | **0.012** |
| Door to needle | 63 (40-80) | 63.5 (IQT= 34.7 – 74.7) | 63 (IQT=45 - 85) | 0.413 |
| Door to groin | 159.5 (IQT= 129.7-207) min | 120 (IQT=103-133.5) min | 165 (IQT = 156-271) min | **0.038\*** |
| Door to Imaging choice | 8 (IQT= 5 – 13) min | 7 (IQT=5-12) min | 9 (IQT=3-13) min | 0.938 |
| Door to therapy decision | 47 (IQT=27.7 – 86.2) min | 40 (IQT=25-65) min | 70 (IQT=32-97) min | 0.061 |
| <u>Imaging to decision</u> | 14 (IQT= 5-28.7) min | 6 (IQT=5-16) min | 22 (IQT=10-45) min | **0.003** |
| <u>Adherence</u> | 41 (85.4%) | 24 (96%) | 17 (73.9%) | **0.030** |
| - Choose of diagnosis | 43 (89.6%) | 24 (96%) | 19 (82.6%) | 0.180 |
| - Choose of therapy | 45 (93.8%) | 25 (100%) | 20 (86.9%) | 0.062 |

